# Ethical reporting of research on violence against women and children during COVID-19: Analysis of 75 studies and recommendations for future guidelines

**DOI:** 10.1101/2022.11.15.22282363

**Authors:** Amber Peterman, Karen Devries, Alessandra Guedes, Joht Singh Chandan, Sonica Minhas, Rachel Qian Hui Lim, Floriza Gennari, Amiya Bhatia

## Abstract

Changes in research practice during the COVID-19 pandemic necessitates renewed attention to ethical protocols and reporting for data collection on sensitive topics. We systematically searched journal publications from the start of the pandemic to November 2021, identifying 75 studies that collected primary data on violence against women and children. We assess the transparency of ethics reporting and adherence to relevant guidelines against a 14-item checklist of best practices. Studies reported adhering to best practices on 31% of scored items with highest reporting for ethical clearance (87%) and informed consent/assent (84/83%) and lowest reporting for facilitating referrals for minors and soliciting participant feedback (both 0%). Violence studies of primary data collected during COVID-19 report on few ethical standards, obscuring stakeholder ability to enforce a ‘do no harm’ approach and to assess the reliability of findings. We offer recommendations and guidelines to improve future reporting and implementation of ethics within violence studies.

## Introduction

Research has demonstrated increases in violence against women and children (VAW/C) across numerous settings during the COVID-19 pandemic.^1–4^ This widespread evidence within a relatively short time period is due to creative use of available administrative data, analysis of ongoing and new data collection efforts. In many parts of the world, data collection during the pandemic required adopting remote or other novel methods to successfully and safely reach and interview participants. Such methods were rarely used for VAW/C prior to the pandemic, particularly in low-income settings.^5^ These efforts challenged teams to ensure the appropriate adaptation of violence-specific safeguarding and ethical protocols. For example, data collected online or over the phone may leave participants vulnerable to lack of privacy, where responses could be overheard or where questionnaire forms or information might be viewed online by perpetrators or household members.^6,7^ In addition, shut-downs and reductions in service provision of violence and social services added complications, particularly for assuring the quality of, and continuous access to, referral services and for implementing response measures for adverse events.^8^ Research teams were forced to choose between collecting violence data in ethically challenging and uncertain contexts or opting to forgo primary data collection altogether. There remains differing opinions as to if, and how, data on VAW/C can be safely and ethically collected in such circumstances. Some early guidance during the pandemic suggested not to collect remote data at all, with the World Health Organization (WHO) and UN Women emphasizing the mantra “*Do not prioritize data over women’s safety.”* ^9^ Others suggested conditions which must be met in order to justify proceeding, including the ability to address safety concerns for participants, implement quality referrals and the critical use of data for immediate policy action.^6,7^ To date, no universal protocols exist for the design and reporting of remote research on VAW/C and ethical review boards are often ill-equipped to advise on violence-specific protocols even in face-to-face data collection efforts. Therefore, the decision of what VAW/C measures to collect and how go about setting up sufficient safeguards during COVID-19 was often made on a case-by-case basis by individual research teams.

This paper provides an initial examination of reporting on ethics and safeguarding within primary data on VAW/C collected during the pandemic. We argue for greater attention to the development, implementation, and reporting of ethics protocols within future studies and publications, both to ensure studies are meeting the commitment to protection participant and researcher safety, and to enhance data quality. To that end, we offer recommendations for researchers and journals across disciplines on which aspects are critical to ensure transparency, offering a 14-item checklist both to guide the study and inform the reader. Although our study presents new findings explicitly addressing challenges and considerations for data collection during COVID-19, poor reporting on ethical practices pre-dates the COVID-19 pandemic.^10^ The stocktaking on ethics for VAW/C research comes at a critical time, when changes in data collection methodologies, advances in information technology and macro-changes across settings have raised debates around harmful practices in data collection. Results suggest the need for higher consensus, guidance, and accountability in order to ensure a ‘do no harm’ approach.

## Methods

### Information sources and search strategy

We searched the studies compiled in the Global Tracker of Studies of VAW/C during COVID-19 (referred to as “the tracker”), compiled from Google scholar and updated weekly starting in April 2020 by the lead author (search terms: “COVID-19” and “violence”).^11^ On November 5^th^, 2021 there were 279 studies in the tracker representing a universe of 3,250 hits on Google scholar. Titles and abstracts were screened by the lead author and all studies including analysis of VAW/C measures during COVID-19 were incorporated in the tracker, including physical, sexual and emotional violence and proxy measures.

### Selection process and inclusion criteria

From the tracker, we selected all peer-reviewed studies where primary data collection methods were used to collect data on VAW/C. The following types of studies were excluded: 1) those in non-English languages, 2) published in grey literature, 3) analysis of administrative or social media data, 3) modeling studies using pre-pandemic data 4) studies analyzing proxy measures of violence (e.g. conflict, attitudes, perceptions of violence risk) and 5) data from services providers or health care workers. Figure A1 provides additional detail on the sample selection.

### Development of criteria for reporting violence research

We developed a checklist for the ethical reporting of violence research drawing on best practice guidelines for implementation of safe data collection for VAW/C established prior to the pandemic.^12–15^ In addition, as the pandemic increased use of remote data collection methods and challenges in accessing service provision, existing guidelines were augmented by key publications outlining best practices for VAW/C research during the pandemic.^6,16^ Lastly, a review of literature was undertaken to explore any studies summarizing or proposing guidelines for ethical reporting of interpersonal violence pre-pandemic, as to build on or complement existing reporting guidelines.^10,17,18^

We developed a 14-item check list of best practices for reporting violence research grouped into four domains: 1) Institutional Review Board (IRB) approval, 2) interviewer selection, training and support, 3) sampling and engaging with respondents, and 4) referrals and adverse events (Table 1). Recognizing that guidelines for the ethical reporting of violence research do not currently exist, checklist items were defined to give studies maximum flexibility for a “yes” coding. For example, for item one regarding IRB approval, a “yes” coding was given regardless of where the IRB was located, or the quality of the IRB assessment. For item two regarding appropriate interviewer selection, any relevant selection criteria was accepted with justification (e.g. prior experience with sensitive topics, sex of interviewer etc.), rather than imposing pre-specified criteria which might differ by setting, survey objectives or target population. For several items, not all studies qualified to be assessed and these were coded as ‘not applicable.’ For example, interviewer selection, training and support items were not applicable for self-administered web-based studies and items 7, 13 and 14, were only relevant to studies focused on collecting VAC data, either from minors or from other adults.

**Table 1.**
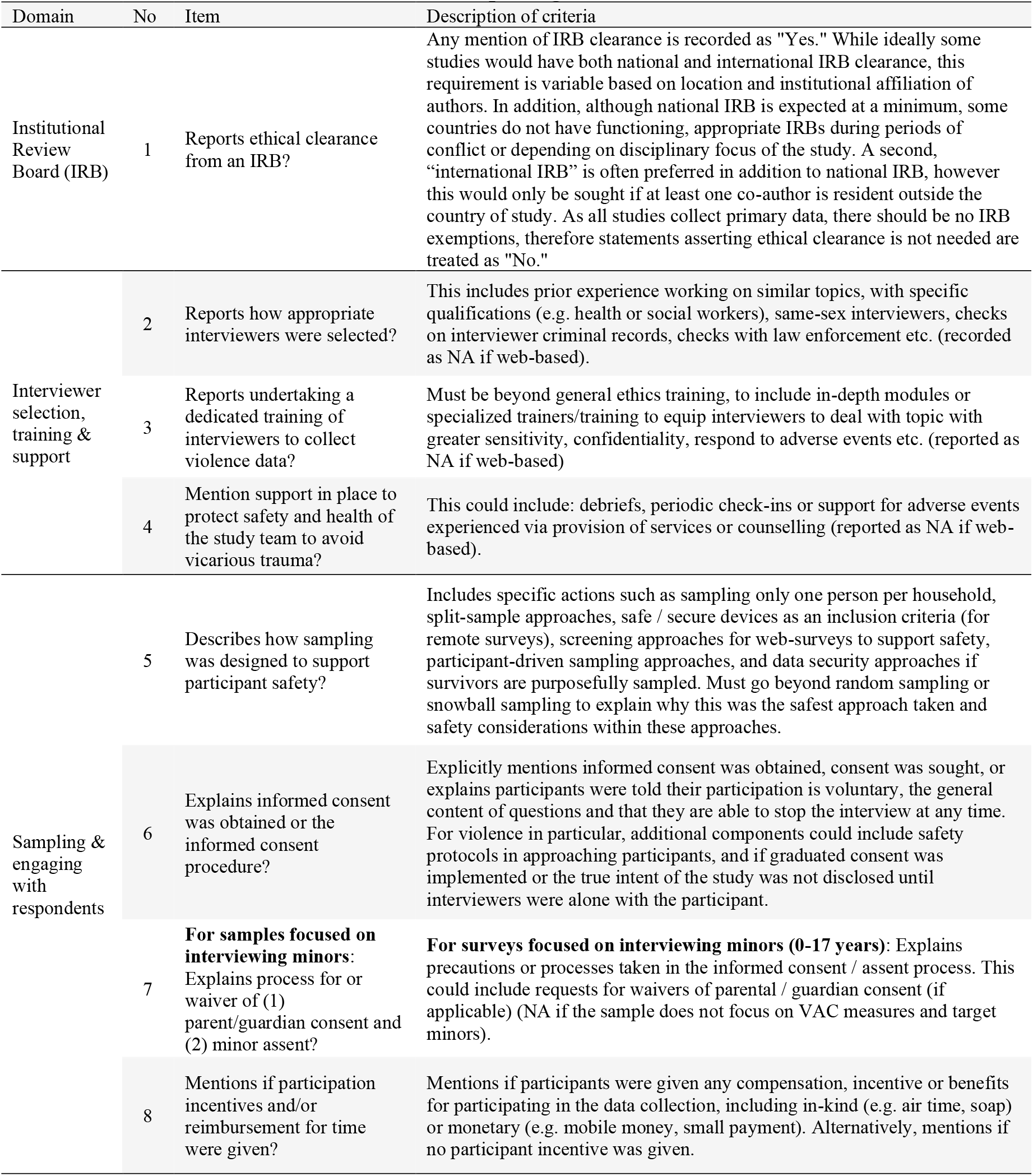
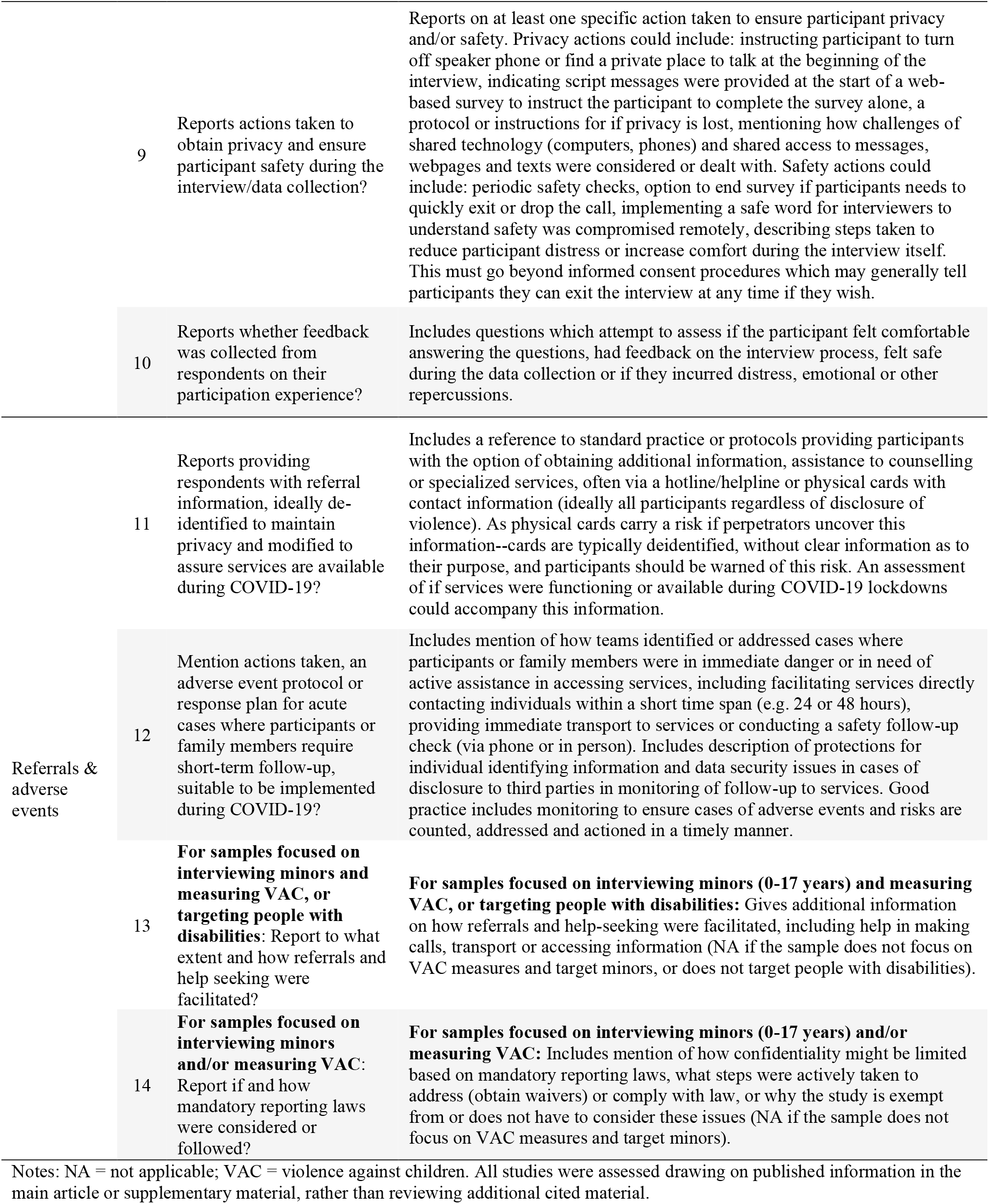
Domain and item definition for ethical reporting.

### Data extraction and analysis

The lead author extracted the background characteristics of each study, including the country of data collection, methodology, mode of data collection and violence measures collected, which was cross-checked by individual reviewers (Table A1). The 14-item checklist was then applied to each study, drawing on information in the main article or supplementary material. To ensure consistency in coding, four reviewers (AP, AB, SM, RL) first used the checklist to score five studies independently and discussed concordance of answers. Subsequently, each study was randomly assigned to two reviewers and scored independently. Considering all studies and all items, the total percentage of discordant results after the first round of scoring was low (4%). Discrepancies were subsequently discussed and resolved, when required, by a third reviewer. Scores for each checklist item were descriptively summarized overall and by study characteristic. In addition, a summary measure was created by averaging the proportion of items reported on (coded as ‘yes’), among the total applicable number of items (all items coded as ‘not appliable’ were not included in this score). Checklist items and summary scores are described overall, by methodology, violence and reporting type, and by mode of data. All descriptive analysis was conducted in Stata version 15.^19^ This study is exempt from ethical approval, as it uses data fully in the public domain and does not use data on human subjects.

## Results

### Studies included

Table 2 describes the adherence to each checklist item among all 75 eligible studies. Most studies collected quantitative data (88%, n=66), in comparison to qualitative data (17%, n=13). The sample was similarly heavily skewed towards collection of VAW data (88%, n=67) and self-reported experience measures (75%, n=64), as compared to VAC data (17%, n=13) or proxy reports (e.g. reporting by household members of violence experienced by children in the same household) (21%, n=16). Web-based methods were the most frequently used (65%, n=49), followed by telephone (17%, n=13) and face-to-face data collection (19%, n=14). The majority of publications were published in public health journals (55%, n=41), while a smaller percentage was in medical journals and other social science journals (23%, n=17 for both disciplines). Data collection occurred in the following regions: South Asia (n=15), Sub-Saharan Africa, Middle East and North Africa, Europe (all n=13), North America (n=12), Asia-Pacific (n=5) and Latin America and the Caribbean (n=3).

**Table 2.** Descriptive statistics for ethical domains and items by study characteristic.

|  | Methodology |  |  |  |  |  | Type of violence |  |  |  | Type of reporting |  |  |  | Modality of data collection |  |  |  |  |  |
| --- | --- | --- | --- | --- | --- | --- | --- | --- | --- | --- | --- | --- | --- | --- | --- | --- | --- | --- | --- | --- |
|  | All studies<br>N=75 |  | Quantitative<br>N=66 |  | Qualitative<br>N=13 |  | VAW<br>N=67 |  | VAC<br>N=13 |  | Self-reports<br>N=64 |  | Proxy reports<br>N=16 |  | Face-to-face<br>N=14 |  | Telephone<br>N=13 |  | Web-based<br>N=49 |  |
|  | n | % | n | % | n | % | n | % | n | % | n | % | n | % | n | % | n | % | n | % |
| *Domain 1: Institutional Review Board* |  |  |  |  |  |  |  |  |  |  |  |  |  |  |  |  |  |  |  |  |
| 1. Ethics clearance | 75 | 0.87 | 66 | 0.88 | 13 | 0.85 | 67 | 0.87 | 13 | 0.92 | 64 | 0.88 | 16 | 0.88 | 14 | 0.71 | 13 | 1.00 | 49 | 0.88 |
| *Domain 2: Interviewer selection, training & support* |  |  |  |  |  |  |  |  |  |  |  |  |  |  |  |  |  |  |  |  |
| 2. Interviewer selection | 30 | 0.33 | 21 | 0.43 | 12 | 0.17 | 29 | 0.34 | 3 | 0.33 | 29 | 0.34 | 3 | 0.33 | 14 | 0.29 | 13 | 0.46 | 4 | 0.00 |
| 3. Interviewer training | 30 | 0.13 | 21 | 0.14 | 12 | 0.08 | 29 | 0.14 | 3 | 0.00 | 29 | 0.14 | 3 | 0.00 | 14 | 0.14 | 13 | 0.15 | 4 | 0.00 |
| 4. Interviewer safety & support | 30 | 0.03 | 21 | 0.00 | 12 | 0.08 | 29 | 0.03 | 3 | 0.00 | 29 | 0.03 | 3 | 0.00 | 14 | 0.00 | 13 | 0.08 | 4 | 0.00 |
| *Domain 3: Sampling & engaging with respondents* |  |  |  |  |  |  |  |  |  |  |  |  |  |  |  |  |  |  |  |  |
| 5. Sampling design | 75 | 0.05 | 66 | 0.03 | 13 | 0.15 | 67 | 0.04 | 13 | 0.08 | 64 | 0.06 | 16 | 0.00 | 14 | 0.00 | 13 | 0.08 | 49 | 0.06 |
| 6. Informed consent | 75 | 0.84 | 66 | 0.85 | 13 | 0.85 | 67 | 0.84 | 13 | 0.85 | 64 | 0.86 | 16 | 0.81 | 14 | 0.79 | 13 | 0.85 | 49 | 0.84 |
| 7. Informed assent (minors) | 6 | 0.83 | 6 | 0.83 | 2 | 1.00 | 3 | 1.00 | 5 | 0.80 | 5 | 0.80 | 2 | 1.00 | 1 | 0.00 | 1 | 1.00 | 4 | 1.00 |
| 8. Participant incentives | 75 | 0.31 | 66 | 0.33 | 13 | 0.23 | 67 | 0.30 | 13 | 0.38 | 64 | 0.28 | 16 | 0.56 | 14 | 0.00 | 13 | 0.31 | 49 | 0.39 |
| 9. Interview privacy & safety | 75 | 0.21 | 66 | 0.21 | 13 | 0.23 | 67 | 0.24 | 13 | 0.15 | 64 | 0.25 | 16 | 0.06 | 14 | 0.21 | 13 | 0.54 | 49 | 0.12 |
| *Domain 4: Referrals & adverse events* |  |  |  |  |  |  |  |  |  |  |  |  |  |  |  |  |  |  |  |  |
| 10. Participant feedback | 75 | 0.00 | 66 | 0.00 | 13 | 0.00 | 67 | 0.00 | 13 | 0.00 | 64 | 0.00 | 16 | 0.00 | 14 | 0.00 | 13 | 0.00 | 49 | 0.00 |
| 11. Referral information | 75 | 0.25 | 66 | 0.27 | 13 | 0.15 | 67 | 0.28 | 13 | 0.15 | 64 | 0.30 | 16 | 0.13 | 14 | 0.21 | 13 | 0.46 | 49 | 0.20 |
| 12. Adverse event protocol | 75 | 0.08 | 66 | 0.08 | 13 | 0.08 | 67 | 0.09 | 13 | 0.00 | 64 | 0.09 | 16 | 0.00 | 14 | 0.14 | 13 | 0.23 | 49 | 0.02 |
| 13. Facilitated referrals (minors) | 6 | 0.00 | 6 | 0.00 | 2 | 0.00 | 3 | 0.00 | 5 | 0.00 | 5 | 0.00 | 2 | 0.00 | 1 | 0.00 | 1 | 0.00 | 4 | 0.00 |
| 14. Mandatory reporting (minors) | 13 | 0.08 | 13 | 0.08 | 2 | 0.00 | 6 | 0.17 | 12 | 0.08 | 7 | 0.00 | 9 | 0.11 | 1 | 0.00 | 2 | 0.00 | 10 | 0.10 |
| Total (among non-missing items) | 75 | 0.31 | 66 | 0.31 | 13 | 0.26 | 67 | 0.31 | 13 | 0.29 | 64 | 0.32 | 16 | 0.29 | 14 | 0.23 | 13 | 0.37 | 49 | 0.31 |

Notes: VAC = violence against children; VAW = Violence against women. Items Q7, Q13 and Q14 only apply to certain studies, those that either target minors for interviews, ask minors violence questions directly or ask about VAC. Items Q2, Q3 and Q4 only apply to studies that use interviews to collect data, and do not apply to web-based data collection. All studies are included in sub-group calculations if they apply, and therefore may appear in more than one category -- e.g. collect both VAW and VAC measures, or collection both qualitative and quantitative data.

### Ethical reporting

Results show adherence to best practices was reported on average for 31% of scored items across the 75 studies. Across the sample, reporting was highest for: ethical clearance (87%) and informed consent/assent (84%/83%). Reporting was lowest for facilitating referrals for minors (0%), soliciting participant feedback (0%), measures to promote interviewer safety and support (3%), safe sampling designs (5%), implementation of adverse event protocols and if mandatory reporting for violence against minors was considered (both at 8%). Other individual items were scored as follows: 33% of studies noted how interviewers were selected to support participant safety, 31% of studies report if incentives were given for participation in the study, 25% of studies report giving some type of violence referral information, 21% report any measure taken to support participant safety and privacy during the interview and 13% report specialized enumerator training on violence topics. Findings suggest little overall variation on the proportion of items reported on by study methodology, type of violence, and type of reporting (questions about self experience of violence versus proxy reporting)—however there is some divergence by modality of data collection. In particular, studies using face-to-face data collection appeared to report fewer items (23% of items), while telephone-based surveys report higher adherence to ethics (37% of items). Finally, we examine ethics reporting by discipline of the journal where studies were published, finding little variation across public health, medical and other social science journals (Table A2). Tables with study-specific results by item are provided in Table A3. Examples of best practice reporting by domain and item are provided using excerpts from the highest scoring papers in Table A4.^20–27^

## Discussion

Our results indicate insufficient overall reporting on ethics of VAW/C research across disciplines. Given the number of studies that fail to report checklist items, our findings raise important questions about the guidance on VAW/C data collection issued by IRBs and the criteria for ethical reporting used and enforced by journals. However, poor reporting on ethical practices also pre-dates the COVID-19 pandemic, suggesting the limited reporting of research ethics we document is illustrative of a larger and more systemic limitation in the field of violence research. For example, a review of studies on childhood sexual abuse in India in 2018 found that only 2/3 of the 51 included studies reported approval by an ethics committee, obtaining informed consent and ensuring confidentiality for participants. Engagement with safeguarding of participants was also poor, with only 25% assessing further risk of sexual abuse and providing services, and no studies describing whether they adhered to the mandatory reporting requirement in India.^10^ In addition, a review of methodology and ethics in 21 studies including gender-based violence outcomes using remote data collection methods (focused on humanitarian and fragile settings) showed only four studies reported offering referral services and only five studies reported any other safety-related measures.^28^ This lack of documentation on adherence to ethical guidance for VAW/C research raises serious concerns about the possibility of harm to research participants and quality of data: we contend that limited attention to ethics affects both participants who disclose violence, what happens when these disclosures are received, as well as the propensity for participants to disclose in the first instance.

Nonetheless, it is possible that both journal editors and ethics committees themselves were affected by COVID-19. For example, a study of Italian ethics committees found that the workload of committees in highly affected areas of the country increased substantially during COVID-19. This, coupled with a decrease in the ability of committee members to work, led some participants to report that “*it was impossible to perform an accurate analysis of the submitted documentation”*^29^ The reprogramming of research to use remote methods required ethics committees and other research stakeholders to rapidly make decisions about new methodologies without centralized guidance. Deviations from established ethical protocols are not unprecedented, and have been deemed acceptable in some circumstances in the context of rapidly evolving humanitarian and emergency situations.^30^ Nonetheless, a review of studies with human participants during COVID-19 found that even more basic ethical reporting has been insufficient—finding up to 24% of observational studies did not report approval by an ethics committee, and up to 38% did not report informed consent from participants.^31^ Our findings suggest that violence research during the pandemic faces similar shortcomings.

Our study has limitations. In general, we were only able to score whether studies mentioned the presence of a particular criterion, rather than on the quality of their adherence to it. In addition, we do not exclude the possibility that studies employed good ethical practices in data collection, without reporting this explicitly in the resulting publication. Finally, there are other ethical aspects not scored here which are also relevant. These include: more general data protection protocols (particularly with technology-facilitated data collection via Apps or interactive voice recall), whether results are actionable and useful to communities, an emphasis on equity and inclusion in sampling, positionality of researchers and whether community members, young people, and survivors were included in the research design and in study steering committees. We choose not to score these criteria, as many of these aspects fall outside the timelines of journal articles or are less likely to be documented in publications.

## Conclusions and Implications

Poor reporting of ethical practices is widespread. In VAW/C research, there is a clear risk of harm to participants if guidance is not followed as well as an impact on the quality of the data produced. In addition, qualitative studies of study interviewers show that they often bear the psychological burden/experience secondary trauma if robust procedures to ensure both their own, and participant, safety are not in place.^32–34^ Our findings point to the importance of the development and use of reporting guidelines for research on VAW/C. Based on our work, the dimensions outlined in Table 1 provide a starting point for such guidance. For violence researchers, dimensions can serve as a checklist, providing strategies that can be incorporated into the design and implementation of research studies. Both ethics committees and journal editors can assess violence research against reporting guidance, similar to the CONSORT or STROBE guidance for reporting of trials and observational studies, respectively. ^35,36^ Additionally, funders could use the checklist to assess research proposals to ensure mechanisms for safety referrals and feedback are integrated into the study from its design. Finally, the checklist could be integrated into efforts to build capacity, particularly in the context of training researchers and data collection teams globally. Efforts to improve the reporting of VAW/C research are an important step to improve the quality and safety of violence research and fulfill the commitments to listen to and learn from participants while ensuring a no harm approach.

## Data Availability

All data produced in the present work are contained in the manuscript

## Authors contributions

Conceptualization (AP, KD, AG, JSC, AB), data curation (AP, SM, RQHL, AB), formal analysis (AP), methodology (AP, KD, AG, JSC, AB), writing – original draft (AP, KD, JSC, FG, AB), writing – review and editing (all authors). AP, SM, RQHL and AB have accessed and verified the data in the manuscript. All authors approved the final version.

## Ethical approval

As per LSHTM Research Ethics Committee guidelines, this study is exempt from ethical approval as it uses data fully in the public domain and does not include data on human subjects (https://www.lshtm.ac.uk/research/research-governance-integrity/ethics). All analysis was undertaken using information available in existing publications and no additional primary data was collected.

## Supplementary Annex Materials: Figures and tables

**Figure A1.**
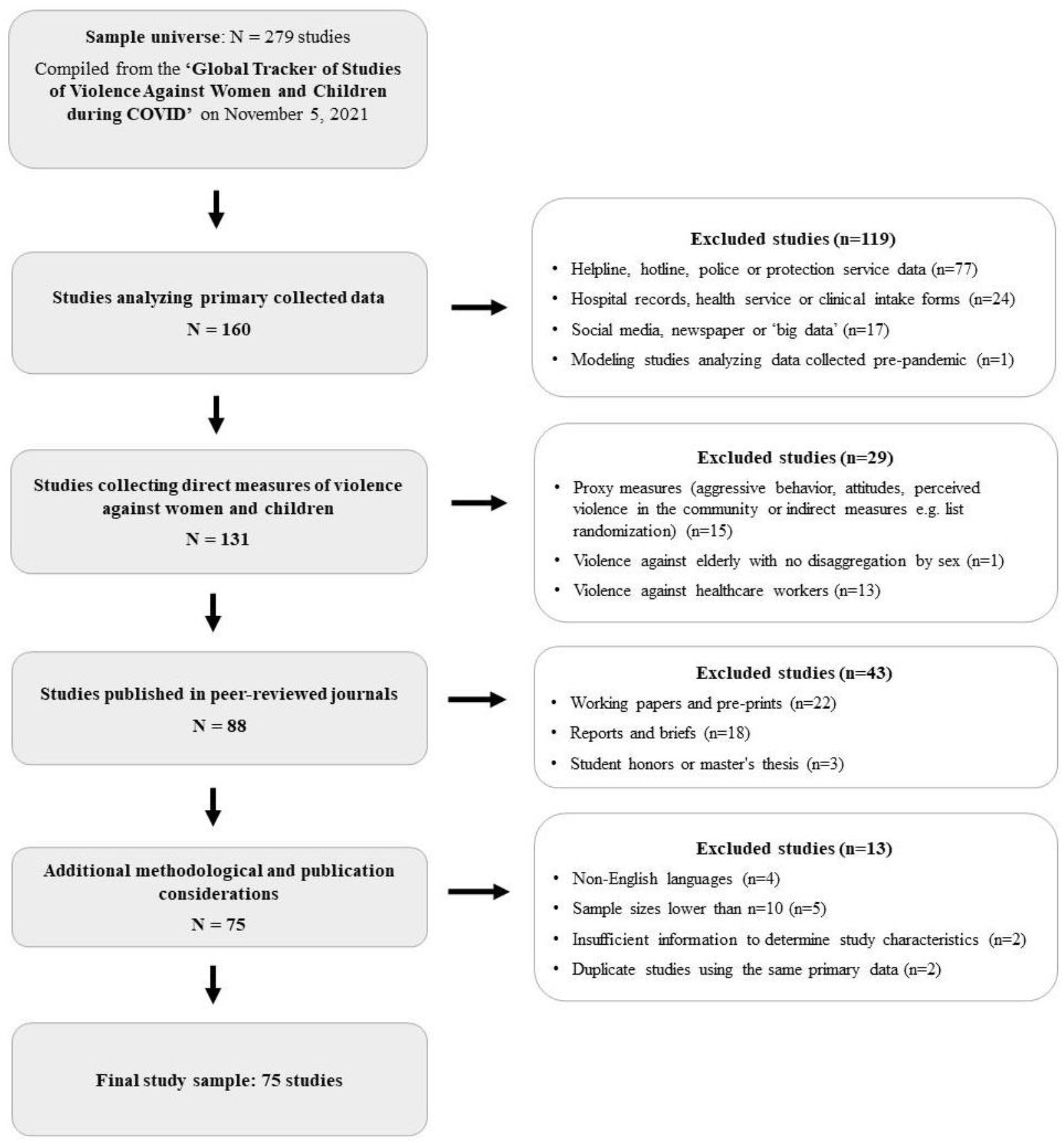
Flow diagram of study selection. Notes: The tracker is an open access database of analysis studies compiled through weekly searches of google scholar (“COVID-19” AND “violence”, hits = 3,250), as well as studies found via multiple listservs, newsletters and social media posts. Parameters for inclusion are: 1) Violence against women and children studies (excludes studies only analyzing violence against men), 2) studies analyzing psychological/emotional, physical and sexual violence experienced in and outside the home, including attitudes and proxy measures (exclude broader forms of gender-based violence, e.g. child marriage, female genital mutilation, child labor etc.) and self-harm (suicide, self-injury) as well as surveys and data from service provider, 3) No restrictions on study methodology, location or type of publication.

**Table A1.**
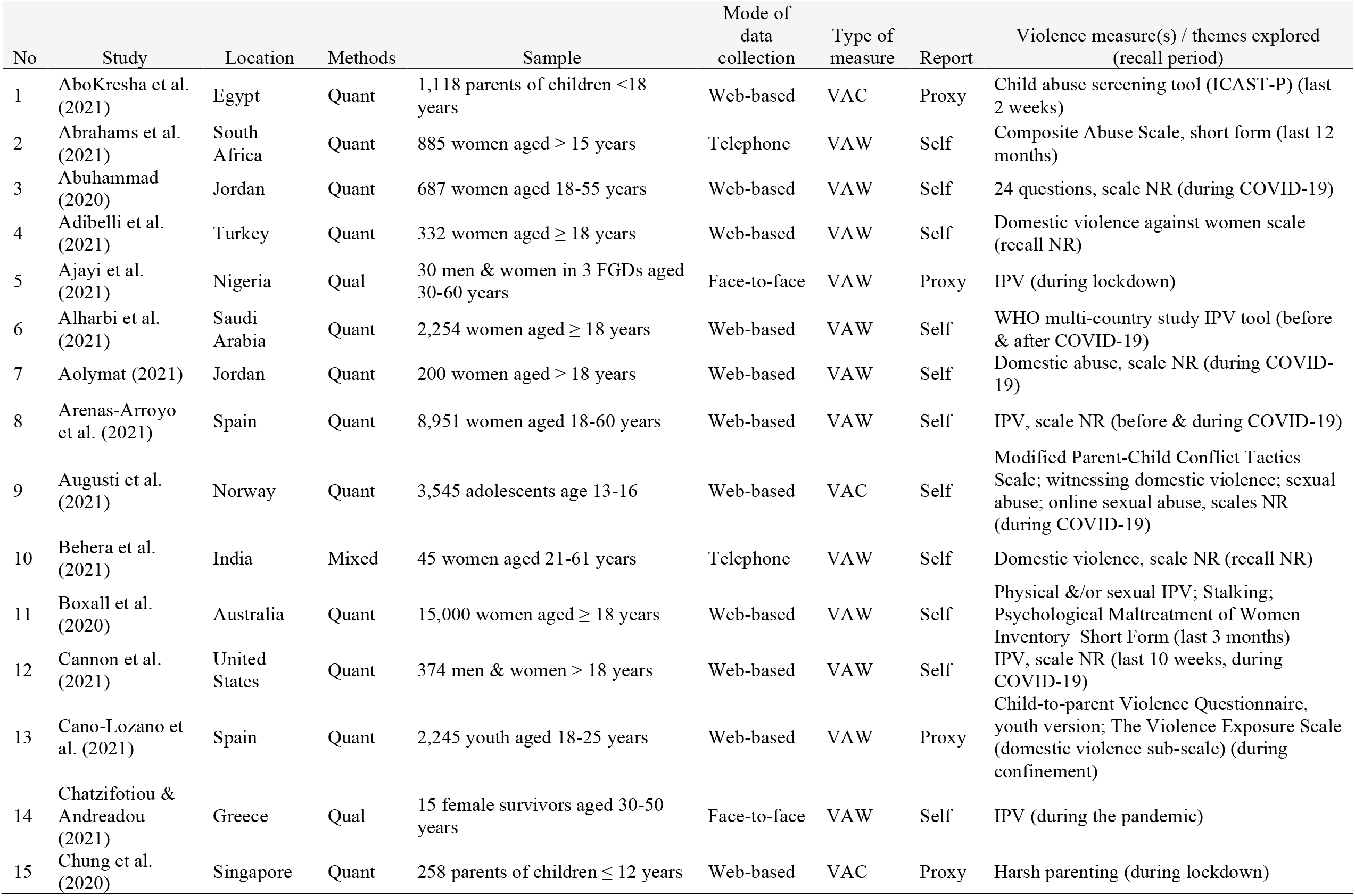
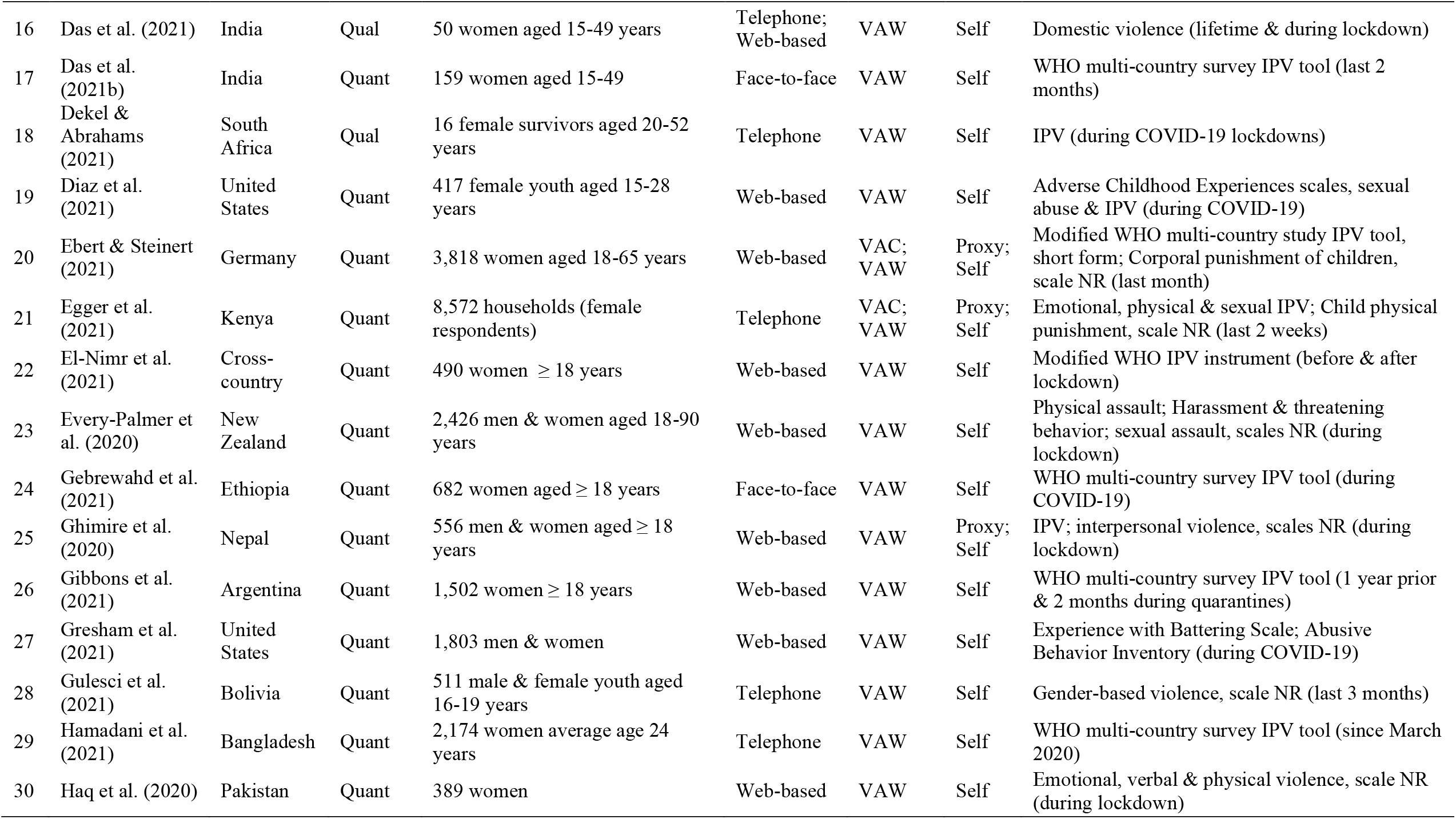
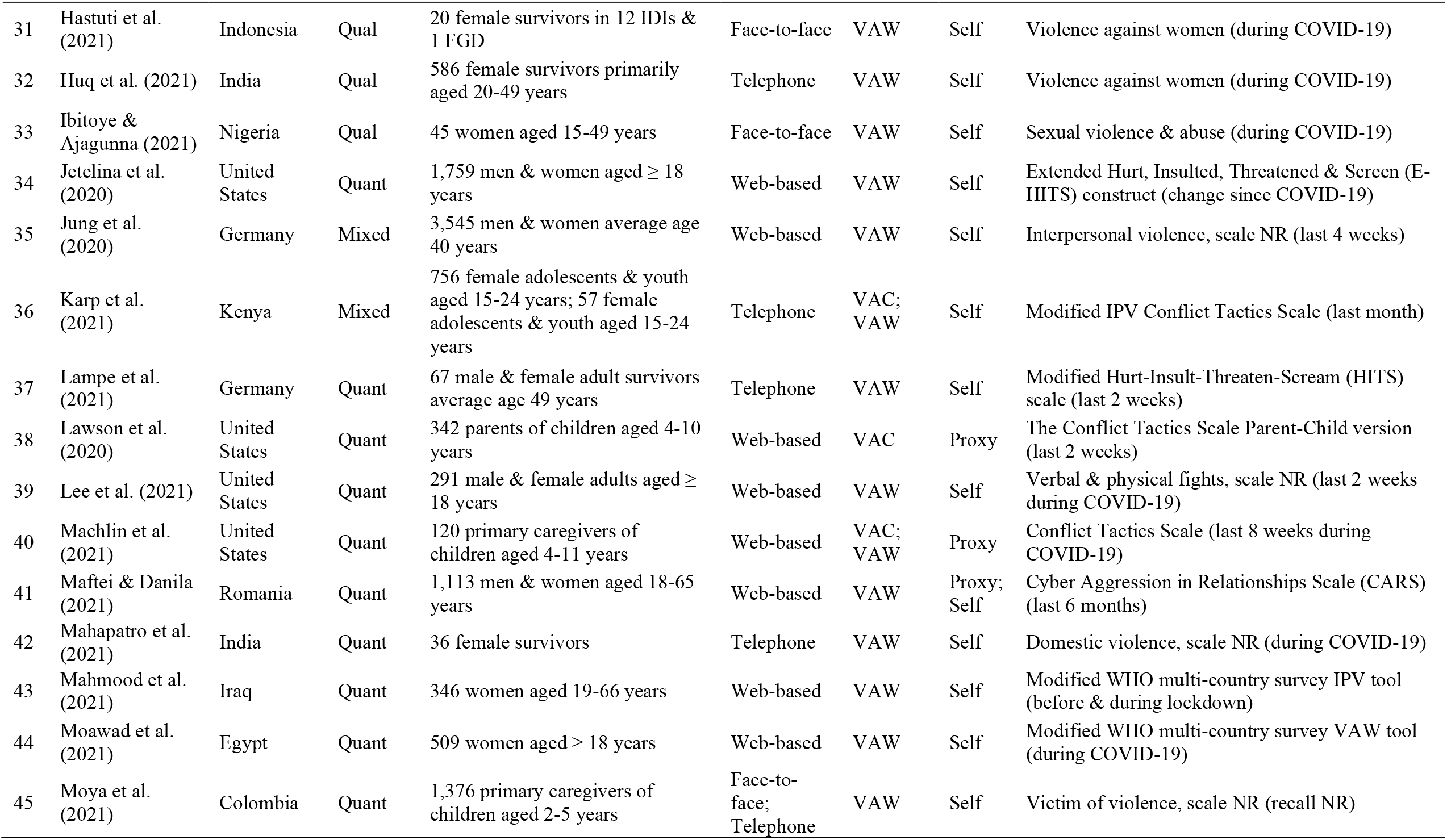
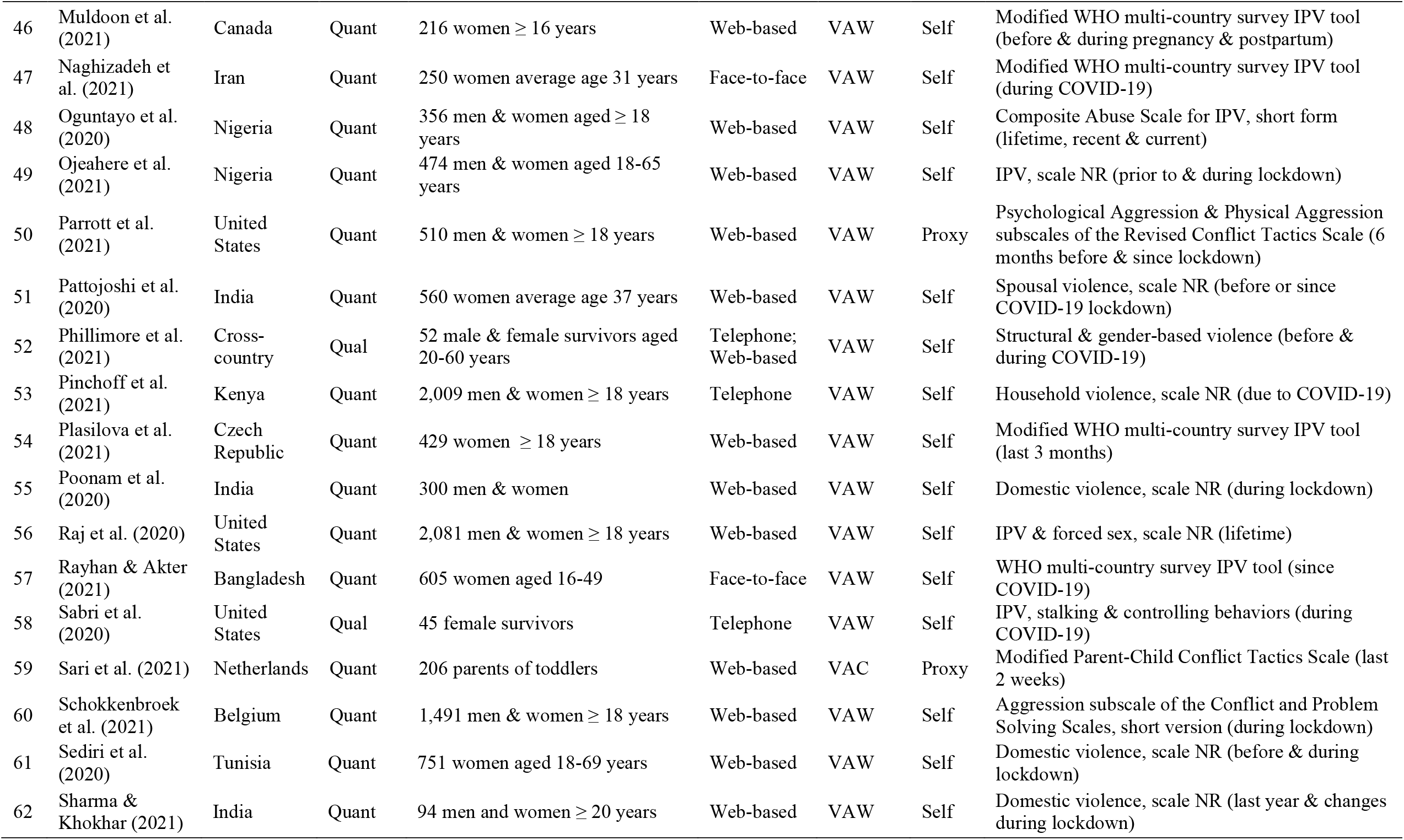
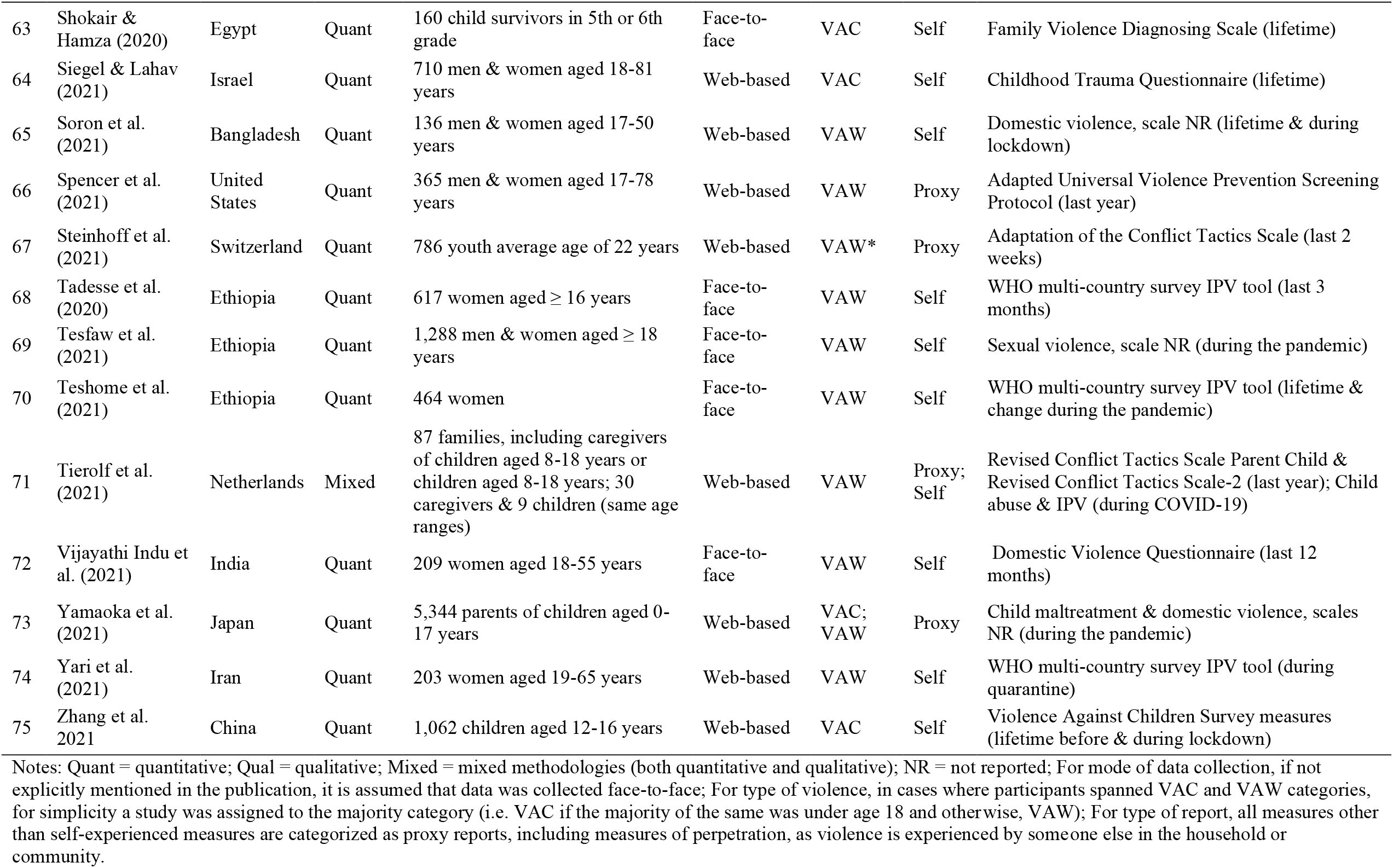
Included study characteristics.

**Table A2.** Descriptive statistics for ethical domains and items by journal discipline of publication.

|  | All studies<br>N=75 |  | Public health<br>N=40 |  | Medical<br>N=17 |  | Social science<br>N=18 |  |
| --- | --- | --- | --- | --- | --- | --- | --- | --- |
| *Domain 1: Institutional Review Board* | n | % | n | % | n | % | n | % |
| 1. Ethics clearance | 75 | 0.87 | 40 | 0.95 | 17 | 0.94 | 18 | 0.61 |
| *Domain 2: Interviewer selection, training & support* |  |  |  |  |  |  |  |  |
| 2. Interviewer selection | 30 | 0.33 | 17 | 0.35 | 5 | 0.40 | 8 | 0.25 |
| 3. Interviewer training | 30 | 0.13 | 17 | 0.12 | 5 | 0.20 | 8 | 0.13 |
| 4. Interviewer safety & support | 30 | 0.03 | 17 | 0.06 | 5 | 0.00 | 8 | 0.00 |
| *Domain 3: Sampling & engaging with respondents* |  |  |  |  |  |  |  |  |
| 5. Sampling design | 75 | 0.05 | 40 | 0.03 | 17 | 0.00 | 18 | 0.17 |
| 6. Informed consent | 75 | 0.84 | 40 | 0.88 | 17 | 0.94 | 18 | 0.67 |
| 7. Informed assent (minors) | 6 | 0.83 | 4 | 1.00 | .. | .. | 2 | 0.50 |
| 8. Participant incentives | 75 | 0.31 | 40 | 0.33 | 17 | 0.35 | 18 | 0.22 |
| 9. Interview privacy & safety | 75 | 0.21 | 40 | 0.23 | 17 | 0.12 | 18 | 0.28 |
| 10. Participant feedback | 75 | 0.00 | 40 | 0.00 | 17 | 0.00 | 18 | 0.00 |
| *Domain 4: Referrals & adverse events* |  |  |  |  |  |  |  |  |
| 11. Referral information | 75 | 0.25 | 40 | 0.30 | 17 | 0.24 | 18 | 0.17 |
| 12. Adverse event protocol | 75 | 0.08 | 40 | 0.13 | 17 | 0.00 | 18 | 0.06 |
| 13. Facilitated referrals (minors) | 6 | 0.00 | 4 | 0.00 | .. | .. | 2 | 0.00 |
| 14. Mandatory reporting (minors) | 13 | 0.08 | 10 | 0.10 | .. | .. | 3 | 0.00 |
| Total (among non-missing items) | 75 | 0.31 | 40 | 0.33 | 17 | 0.31 | 18 | 0.26 |

Notes: VAC = violence against children; VAW = Violence against women. Items Q7, Q13 and Q14 only apply to certain studies, those that either target minors for interviews, ask minors violence questions directly or ask about VAC. Items Q2, Q3 and Q4 only apply to studies that use interviews to collect data, and do not apply to web-based data collection. All studies are included in sub-group calculations if they apply, and therefore may appear in more than one category -- e.g. collect both VAW and VAC measures, or collection both qualitative and quantitative data.

**Table A3.**
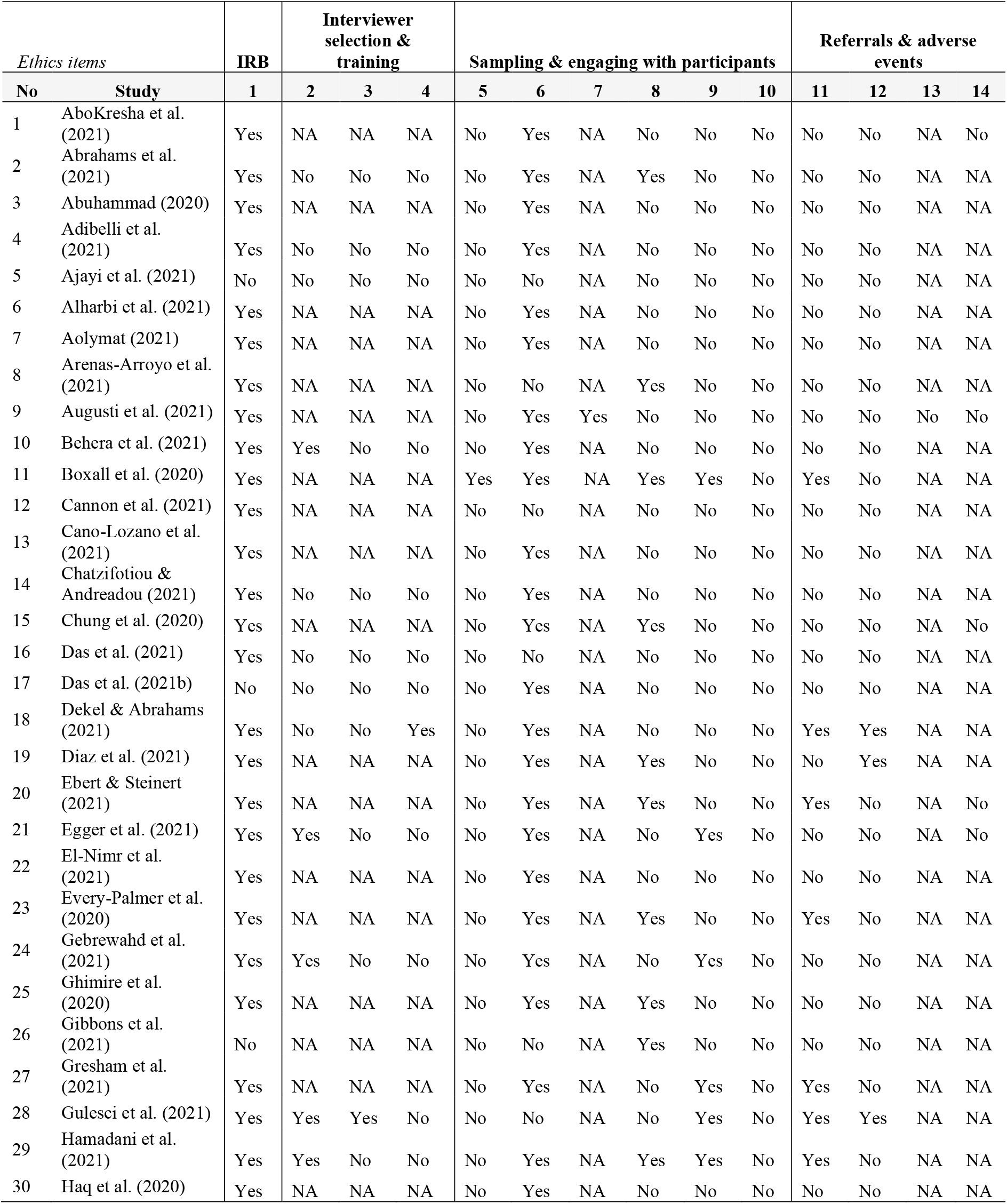
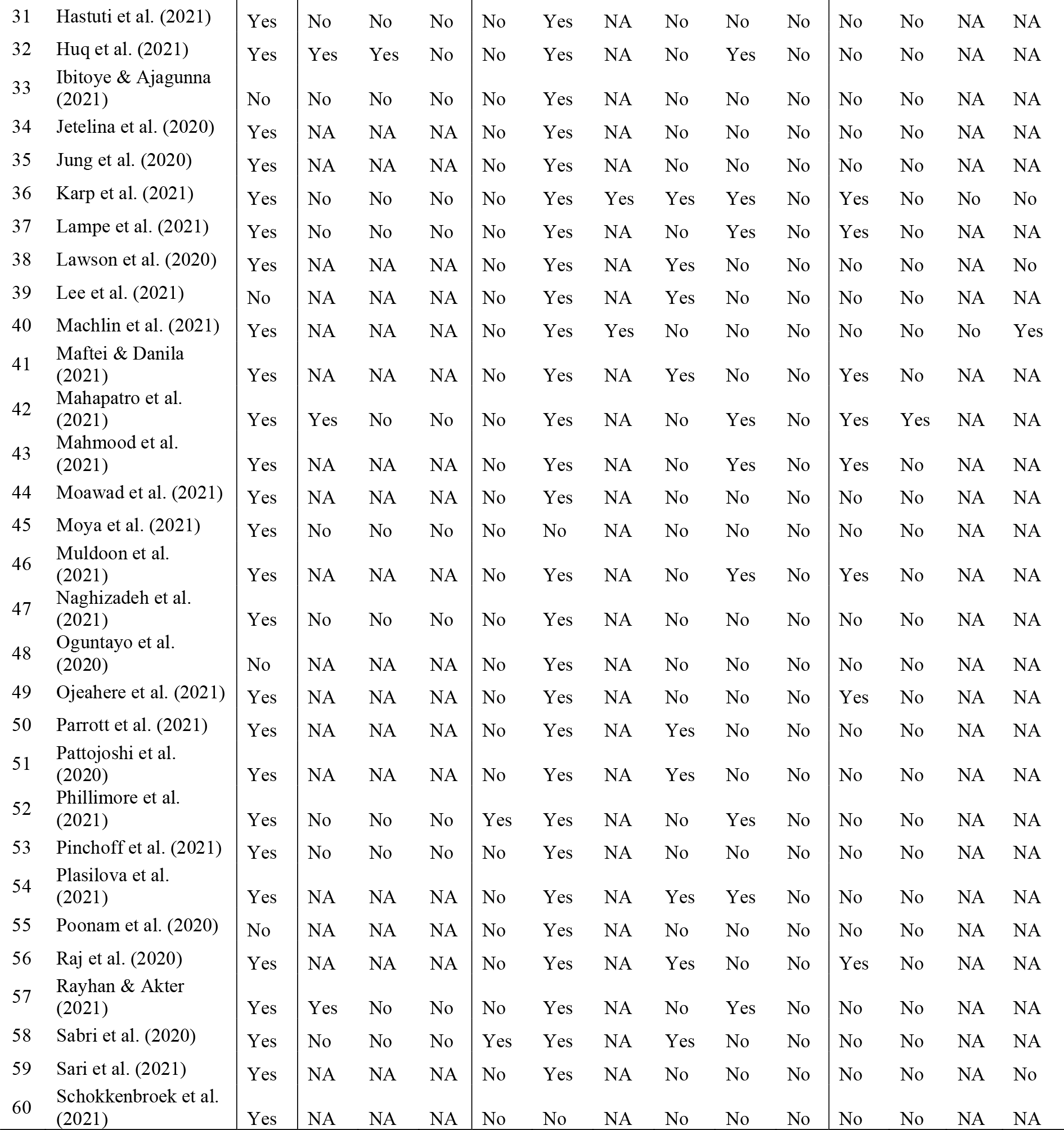
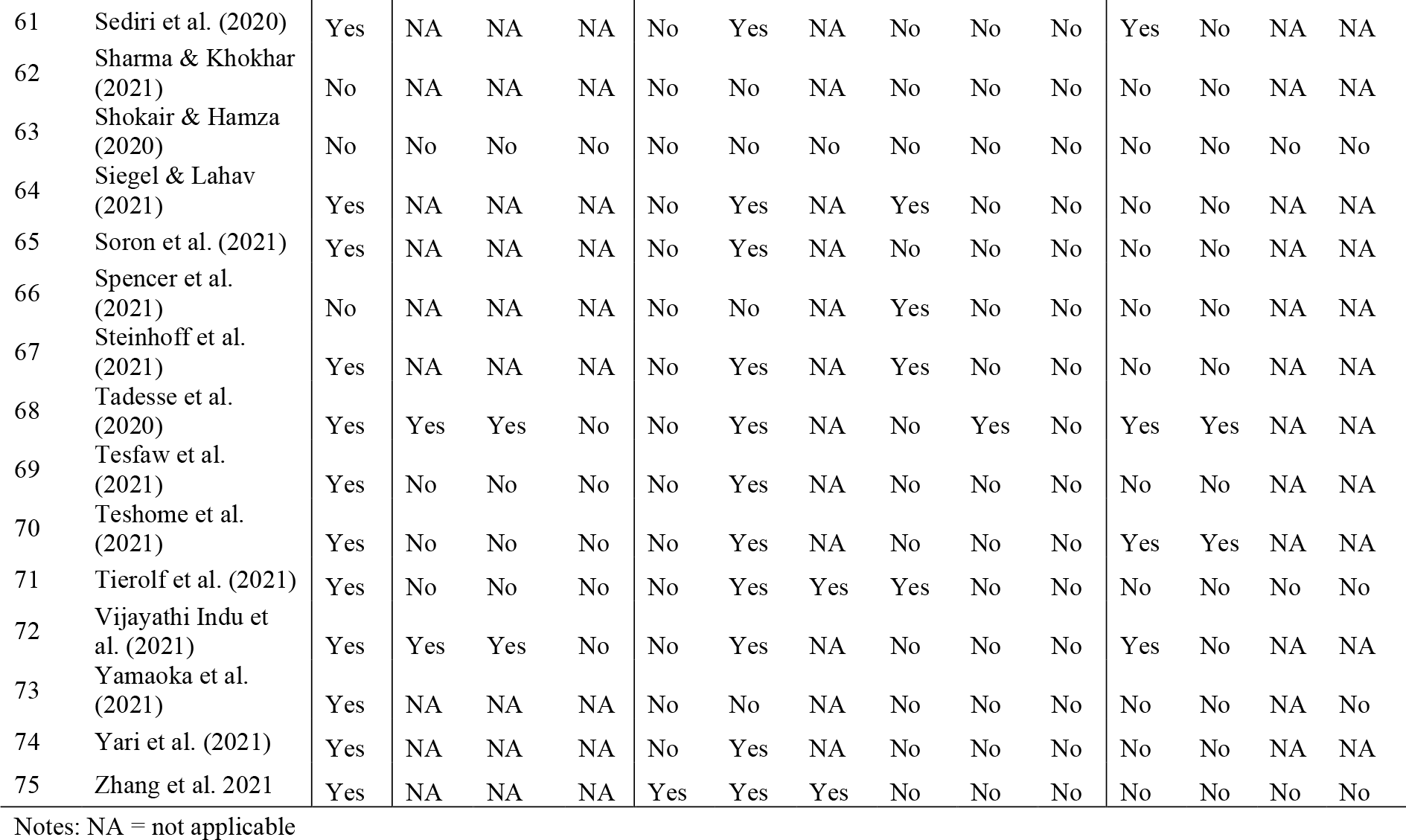
Ethics coding for individual items by study.

**Table A4:** Good practice box on ethical reporting from high scoring studies.

| Domains | Illustrative text from high scoring studies |
| --- | --- |
| **Institutional Review Board (IRB)**<br>[Item 1] | All work was approved by the institutional ethical review boards at the International Center for Diarrhoeal Diseases Research, Bangladesh, and Melbourne Health (2016.269) (Hamadani et al. 2021). [Item 1]<br>Ethical approval was obtained from Dream Science and Technology Institutional Health Research Ethics Review Committee with approval letter of DSTC/ DHS/002/2020. Then, permission letter was written for Dessie city administration office (Tadesse et al. 2020). [Item 1] |
| **Interviewer selection, training & support**<br>[Items 2-4] | To guarantee respondent's safety, enumerators were trained in each case by an expert on Child Safeguarding Policy following stringent ethical guidelines on how to ask these questions. Enumerators were instructed to take measures to verify the privacy of the interviews. Same-sex enumerators were used when possible (Gulesci et al. 2021). [Items 2, 3, 9]<br>Two days training were provided for data collectors and supervisors regarding the sensitivity and personal nature of the questions, objective, and how to approach study participants with ensuring their privacy (Tadesse et al. 2020). [Items 3, 9]<br>We recognized that support for participants was vital and thus, prior to each interview we asked each social worker whether he/she would be willing to meet with the women after the interview, if she felt this was needed. This would have been followed up by the first author to ensure that all participants requesting this service, received it, however, no participants requested additional therapy. Psychological support was also arranged for the first author, who conducted the interviews (Dekel & Abrahams 2021) [Item 4, 12] |
| **Sampling & engaging with respondents**<br>[Items 5-10] | The safety of women participating in the survey was of paramount concern. Given the sensitive nature of the information being collected, a range of safety measures were employed. Safety measures used as part of the survey included:<br>• Potential respondents were approached by a social research company with an established online panel rather than by the AIC because it would be less likely to raise the suspicion of an abusive partner;<br>• The survey was designed with multiple landing pages and eligibility questions (including a 'safety trap') to screen out ineligible participants (eg men) from accessing the survey;<br>• The content of the survey, and its explicit focus on women, was revealed to respondents only after they had gone through multiple landing pages, stated they met the eligibility criteria and confirmed that they were in a safe place where they were not being observed;<br>• Women were advised in the information page that, if they felt that answering questions about their relationship experiences would cause them distress or make them unsafe, they should not complete the survey;<br>• Women who closed the survey at any point were not approached again;<br>• The survey was kept as short as possible and piloted to ensure that women would spend no more than 10 minutes completing all the questions; and<br>• Participants were provided with information about a range of support services, including services that could be contacted online or over the phone. Finally, all of the survey questions were closed-response, meaning that respondents did not have to write any responses. This limited the potential for abusive partners to use keyloggers to access information their partners provided in the survey (Boxall et al. 2020) [Items 5, 9, 11]<br>The women were approached by the female counselors of MSSK over the phone. They had to be telephoned several times before they could be reached. A few limitations that were encountered in virtual communication included fear of a lack of privacy and confidentiality. In many cases, women were reluctant to answer questions about violence that they deemed unimportant in comparison to |
|  | their immediate concerns regarding food, money, health, and the prevailing situation. Some women refused to answer questions over the phone and wanted to talk through a physical confrontation in MSSK only. However, measures were taken to minimize the risk of this non-response bias by allowing respondents to choose a suitable date and time. Thereafter, the counselor tried several times to contact the women when they could respond without the fear of their conversation being interrupted or eavesdropped. This was essential to guarantee their safety, apart from pre-serving the ethics and protocols of research so that respondents were comfortable enough to respond freely (Mahapatro et al. 2021). [Items 5, 9]<br>We had obtained verbal consent from individual study participants before beginning of data collection (Tadesse et al. 2020) [Item 6]<br>Verbal informed consent was obtained from participants aged 18 and older; those younger than 18 provided verbal assent with a parent/guardian providing consent (Karp et al. 2021) [Item 7]<br>Participants were not compensated for their time in this study, although they had been compensated during the main trial at each visit (baseline, midline, endline). Women were warned before commencement of the intimate partner violence module and encouraged to seek privacy; they could decline to answer any module (Hamadani et al. 2021). [Items 8, 9]<br>In the case of phone interviews, additional steps were taken to prevent potential perpetrators from listening to participants' answers. In particular, the interviewer provided examples of what types of actions are considered as violent; participants were asked to answer only "yes" or "no" and given the option of not answering the question if they did not feel comfortable with it. (Gulesci et al. 2021) [Item 9] |
| **Referrals & adverse events**<br>[Items 11-14] | Authors provided text from the questionnaire in a technical appendix: "If you feel upset about anything (now or while completing the survey), the details of someone you can talk to will be made available to you. We have also provided the contact details for services that can support women who are experiencing violence. If you need any kind of help or support, it is available" (Boxall et al. 2020). [Item 11]<br>Respondents were also provided with a list of all the institutions where a violence victim can receive help and protection as well as the procedure to file a complaint (Appendix A.2 shows pictures of the material given to participants). Enumerators received an adverse event protocol explaining what they had to do in cases of abuse (Gulesci et al. 2021). [Items 11, 12]<br>The women who were victims of IPV at the time of data collection were reassured and counseled. However, women who experienced severe IPV and were in need of help were taken to Dessie referral hospital counseling care units (Tadesse et al. 2020). [Item 12]<br>As per the government guidelines, follow-up measures were taken by the counselors and they were expected to call each survivor and understand their situation, extend support, and ensure their safety (Mahapatro et al. 2021). [Item 12]<br>If parents or children reported prior experiences of family violence at baseline, the study reported violence exposure to child protective services if not previously reported (Machlin et al. 2021) [Item 14] |

Notes: Table is based primarily on the five studies which reported on more than half the items in the ethics reporting checklist (Boxall et al. 2020, 75%; Tadesse et al. 2020, 64%; Gulesci et al. 2021, 55%; Hamadani et al. 2021, 55%; Mahapatro et al. 2021, 55%). In addition, examples are augmented by additional studies providing examples of rarely reported items (Dekel & Abrahams 2021; Karp et al. 2021; Machlin et al. 2021). All text included is a direct quotation.

## Works cited

1. Bhatia A, Fabbri C, Cerna-Turoff I, Turner E, Lokot M, Warria A, et al. Violence against children during the COVID-19 pandemic. Bull World Health Organ. 2021 Oct 1;99(10):730–8. doi:10.2471/BLT.20.283051

2. Cappa C, Jijon I. COVID-19 and violence against children: A review of early studies. Child Abuse Negl. 2021 Jun 1;116:105053. doi:10.1016/j.chiabu.2021.105053

3. Flor LS, Friedman J, Spencer CN, Cagney J, Arrieta A, Herbert ME, et al. Quantifying the effects of the COVID-19 pandemic on gender equality on health, social, and economic indicators: a comprehensive review of data from March, 2020, to September, 2021. The Lancet. 2022 Mar 2. doi:10.1016/S0140-6736(22)00008-3

4. Peterman A, Potts A, O’Donnell M, Thompson K, Shah N, Oertelt-Prigione S, van Gelder N. Pandemics and Violence Against Women and Children [Internet]. Washington, DC: Center for Global Development; 2020 Apr 1 [cited 2022 Apr 21]. Available from: https://www.cgdev.org/publication/pandemics-and-violence-against-women-and-children

5. Seff I, Vahedi L, McNelly S, Kormawa E, Stark L. Remote evaluations of violence against women and girls interventions: a rapid scoping review of tools, ethics and safety. BMJ Glob Health. 2021 Sep 1;6(9):e006780. doi:10.1136/bmjgh-2021-006780

6. SVRI. Knowledge Exchange Pivoting to remote research on violence against women during COVID-19 [Internet]. Sexual Violence Research Iniative; 2020. [cited 2022 Apr 7] Available from: https://tinyurl.com/3b8z4bky

7. UNICEF. Research on violence against children during the COVID-19 pandemic: Guidance to inform ethical data collection and evidence generation [Internet]. New York: UNICEF;2020 [cited 2022 Apr 1]. Available from: https://tinyurl.com/yyf52ewb

8. Bhatia A, Fabbri C, Cerna-Turoff I, Tanton C, Knight L, Turner E, et al. COVID-19 response measures and violence against children. Bull World Health Organ. 2020 Sep 1;98(9):583. doi:10.2471/BLT.20.263467

9. UN Women. Violence against women and girls data collection during COVID-19 [Internet]. UN Women – Headquarters. [cited 2022 Apr 7]. Available from: https://tinyurl.com/yckndzfb

10. Dayal R, Kalokhe AS, Choudhry V, Pillai D, Beier K, Patel V. Ethical and definitional considerations in research on child sexual violence in India. BMC Public Health. 2018 Sep 27;18(1):1144. [cited 2022 Apr 7] Available from: doi:10.1186/s12889-018-6036-y

11. Peterman, A. Studies of COVID-19 & Violence Against Women & Children: A Global Tracker.xlsx [Internet]. Google Docs. 2020 [cited 2022 May 27]. Available from: https://tinyurl.com/49k5ez69

12. World Health Organization. Global Programme on Evidence for Health Policy. Putting women first : ethical and safety recommendations for research on domestic violence against women [Internet]. Geneva: World Health Organization; 2001 [cited 2022 Apr 7]. Report No.: WHO/FCH/GWH/01.1. 31p. Available from: https://apps.who.int/iris/handle/10665/65893

13. World Health Organization. Ethical and safety recommendations for intervention research on violence against women: building on lessons from the WHO publication putting women first: ethical and safety recommendations for research on domestic violence against women [Internet]. Geneva: World Health Organization; 2016 [cited 2022 May 30]. 40 p. Available from: https://apps.who.int/iris/handle/10665/251759

14. Centers for Disease Control and Prevention. Critical Elements of Interviewer Training for Engaging Children and Adolescents in Global Violence Research: Best Practices and Lessons Learned from the Violence Against Children Survey [Internet]. Atlanta, GA: National Center for Injury Prevention and Control, Centers for Disease Control and Prevention; 2017 [cited 2022 May 30]. Available from https://tinyurl.com/nxek9rnr

15. UNICEF. Ethical Principles, Dilemmas and Risks in Collecting Data on Violence Against Children [Internet]. New York: UNICEF; 2012 [cited 2022 Apr 7]. 93p. Available from: https://tinyurl.com/37tbmvw4

16. UNICEF. Research on violence against children during the COVID-19 pandemic: Guidance to inform ethical data collection and evidence generation [Internet]. New York: UNICEF; 2020 [cited 2022 Apr 21]. Available from: https://tinyurl.com/yyf52ewb

17. Calia C, Guerra C, Reid C, Marley C, Barrera P, Oshodi AGT, et al. Developing an Evidence-base to Guide Ethical Action in Global Challenges Research in Complex and Fragile Contexts: A Scoping Review of the Literature. Ethics Soc Welf. 2022 Jan 2;16(1):54–72. doi:10.1080/17496535.2021.1916830

18. Orr DMR, Daoust G, Dyvik SL, Puhan SS, Boddy J. Safeguarding in International Development Research: Evidence Review [Internet]. London: UK Collaborative on Development Research; 2019. 63p. [cited 2022 May 30] Available from: https://core.ac.uk/download/pdf/217284902.pdf

19. Stata. Statistical software for data science [Internet]. [cited 2022 May 30]. Available from: https://www.stata.com/

20. Boxall H, Morgan A. Who is most at risk of physical and sexual partner violence and coercive control during the COVID-19 pandemic? [Internet]. Australian Institute of Criminology; 2021 [cited 2022 Mar 1]. Available from: https://www.aic.gov.au/publications/tandi/tandi618

21. Dekel B, Abrahams N. ‘I will rather be killed by corona than by him…’: Experiences of abused women seeking shelter during South Africa’s COVID-19 lockdown. PLOS ONE. 2021 Oct 28;16(10):e0259275. doi: 10.1371/journal.pone.0259275

22. Gulesci S, Puente–Beccar M, Ubfal D. Can youth empowerment programs reduce violence against girls during the COVID-19 pandemic? J Dev Econ. 2021 Nov;153:102716. doi:10.1016/j.jdeveco.2021.102716

23. Hamadani JD, Hasan MI, Baldi AJ, Hossain SJ, Shiraji S, Bhuiyan MSA, et al. Immediate impact of stay-at-home orders to control COVID-19 transmission on socioeconomic conditions, food insecurity, mental health, and intimate partner violence in Bangladeshi women and their families: an interrupted time series. Lancet Glob Health. 2020 Nov;8(11):e1380–9. doi: 10.1016/S2214-109X(20)30366-1

24. Karp C, Moreau C, Sheehy G, Anjur-Dietrich S, Mbushi F, Muluve E, et al. Youth Relationships in the Era of COVID-19: A Mixed-Methods Study Among Adolescent Girls and Young Women in Kenya. J Adolesc Health. 2021 Nov;69(5):754–61. doi:10.1016/j.jadohealth.2021.07.017

25. Machlin L, Gruhn MA, Miller AB, Milojevich HM, Motton S, Findley AM, et al. Predictors of family violence in North Carolina following initial COVID-19 stay-at-home orders. Child Abuse Negl. 2021 Oct;105376. doi: 10.1016/j.chiabu.2021.105376

26. Mahapatro M, Prasad MM, Singh SP. Role of Social Support in Women facing Domestic Violence during Lockdown of Covid-19 while Cohabiting with the Abusers: Analysis of Cases Registered with the Family Counseling Centre, Alwar, India. J Fam Issues. 2021 Nov;42(11):2609–24. doi:10.1177/0192513X20984496

27. Tadesse AW, Tarekegn SM, Wagaw GB, Muluneh MD, Kassa AM. Prevalence and Associated Factors of Intimate Partner Violence Among Married Women During COVID-19 Pandemic Restrictions: A Community-Based Study. J Interpers Violence. 2020 Dec 8;088626052097622. doi:10.1177/0886260520976222

28. Vahedi L, Qushua N, Seff I, Doering M, Stoll C, Bartels SA, et al. Methodological and Ethical Implications of Using Remote Data Collection Tools to Measure Sexual and Reproductive Health and Gender-Based Violence Outcomes among Women and Girls in Humanitarian and Fragile Settings: A Mixed Methods Systematic Review of Peer-Reviewed Research. Trauma Violence Abuse. 2022 May 24;15248380221097440. doi:10.1177/15248380221097439

29. Monaco E, Pisano L, Nasato L, Lorenzoni G, Gregori D, Martinato M. Ethics committees in the time of COVID-19. Epidemiol Prev. 2020 Dec;44(5-6 Suppl 2):113–8. [cited 2022 May 30]. Available from: https://www.ncbi.nlm.nih.gov/pmc/articles/PMC7784177/

30. Richardson T, Johnston AM, Draper H. A Systematic Review of Ebola Treatment Trials to Assess the Extent to Which They Adhere to Ethical Guidelines. PLOS ONE. 2017 Jan 17;12(1):e0168975. doi:10.1371/journal.pone.0168975

31. O’Sullivan L, Killeen RP, Doran P, Crowley RK. Adherence with reporting of ethical standards in COVID-19 human studies: a rapid review. BMC Med Ethics. 2021 Jun 28;22(1):80. doi:10.1186/s12910-021-00649-9

32. Schulz P, Kreft AK, Touquet H, Martin S. Self-care for gender-based violence researchers – Beyond bubble baths and chocolate pralines. Qualitative Research. 2022. doi: 10.1177/14687941221087868

33. Smith AM, Hamilton AB, Loeb T, Pemberton J, Wyatt GE. Reactions of Novice Interviewers Conducting Trauma Research With Marginalized Communities: A Qualitative Analysis. J Interpers Violence. 2021 Nov 1;36(21–22):NP12176–97. doi:10.1177/0886260519889925

34. van der Merwe A, Hunt X. Secondary trauma among trauma researchers: Lessons from the field. Psychol Trauma Theory Res Pract Policy. 2019;11(1):10–8. doi: 10.1037/tra0000414

35. Consort. Consolidated Standards of Reporting Trials [Internet]. [cited 2022 May 25]. Available from: http://www.consort-statement.org/

36. STROBE. Strengthening the reporting of observational studies in epidemiology [Internet]. [cited 2022 May 25]. Available from: https://www.strobe-statement.org/

## References

AboKresha SA, Abdelkreem E, Ali RAE. Impact of COVID-19 pandemic and related isolation measures on violence against children in Egypt. J Egypt Public Health Assoc. 2021;96(1):11. doi:10.1186/s42506-021-00071-4

Abrahams Z, Boisits S, Schneider M, Prince M, Lund C. The relationship between common mental disorders (CMDs), food insecurity and domestic violence in pregnant women during the COVID-19 lockdown in Cape Town, South Africa. Soc Psychiatry Psychiatr Epidemiol. 2022;57(1):37–46. doi:10.1007/s00127-021-02140-7

Abuhammad S. Violence against Jordanian Women during COVID-19 Outbreak. Int J Clin Pract. 2021;75(3). doi:10.1111/ijcp.13824

Adibelli D, Sümen A, Teskereci G. Domestic violence against women during the Covid-19 pandemic: Turkey sample. Health Care for Women International. 2021;42(3):335–350. doi:10.1080/07399332.2021.1885408

Ajayi OA, Ibrahim AT, Kayode OE. COVID-19 Pandemic Lockdown, Intimate Partner Violence and Family Cohesion in Kano, Nigeria. Rev. Universitara Sociologie. 2021:83. [cited 2022 Mar 1]. Available from: https://tinyurl.com/yckzxfh6

Alharbi FF, Alkheraiji MA, Aljumah AA, Al-Eissa M, Qasim SS, Alaqeel MK. Domestic Violence Against Married Women During the COVID-19 Quarantine in Saudi Arabia. Cureus. Published online May 25, 2021. doi:10.7759/cureus.15231

Arenas-Arroyo E, Fernandez-Kranz D, Nollenberger N. Intimate partner violence under forced cohabitation and economic stress: Evidence from the COVID-19 pandemic. Journal of Public Economics. 2021;194:104350. doi:10.1016/j.jpubeco.2020.104350

Augusti EM, Sætren SS, Hafstad GS. Violence and abuse experiences and associated risk factors during the COVID-19 outbreak in a population-based sample of Norwegian adolescents. Child Abuse & Neglect. 2021;118:105156. doi:10.1016/j.chiabu.2021.105156

Behera RR, Borgohain J, Rath CS, Patnaik P. Well-being of female domestic workers during three months of COVID-19 lockdown: Case study from IIT Kharagpur campus. Indian Journal of Health and Wellbeing. 2021 Mar 1;12(1):83–92. [cited 2022 Mar 1]. Available from: https://tinyurl.com/yc6tbh82

Boxall H, Morgan A. Who is most at risk of physical and sexual partner violence and coercive control during the COVID-19 pandemic? [Internet]. Australian Institute of Criminology; 2021 [cited 2022 Mar 1]. Available from: https://www.aic.gov.au/publications/tandi/tandi618

Boxall H, Morgan A, Brown R. The prevalence of domestic violence among women during the COVID-19 pandemic. Australasian Policing. 2020 Sep;12(3):38–46. [cited 2022 Mar 1]. Available from: https://search.informit.org/doi/epdf/10.3316/informit.435862482298266

Cannon CEB, Ferreira R, Buttell F, First J. COVID-19, Intimate Partner Violence, and Communication Ecologies. American Behavioral Scientist. 2021;65(7):992–1013. doi:10.1177/0002764221992826

Cano-Lozano MC, Navas-Martínez MJ, Contreras L. Child-to-Parent Violence during Confinement Due to COVID-19: Relationship with Other Forms of Family Violence and Psychosocial Stressors in Spanish Youth. Sustainability. 2021;13(20):11431. doi:10.3390/su132011431

Chatzifotiou S, Andreadou D. Domestic Violence During the Time of the COVID-19 Pandemic: Experiences and Coping Behavior of Women from Northern Greece. International Perspectives in Psychology. 2021;10(3):180–187. doi:10.1027/2157-3891/a000021

Chung G, Lanier P,Wong PYJ. Mediating Effects of Parental Stress on Harsh Parenting and Parent-Child Relationship during Coronavirus (COVID-19) Pandemic in Singapore. J Fam Viol. 2022;1(37):801–812. doi:10.1007/s10896-020-00200-1

Das et al. Locked in: An Evidence based Study on Domestic Violence during COVID-19 in the Hilly Region of India. Indian Journal of Public Health Research & Development. 2021;12(3):6–15. doi:10.37506/ijphrd.v12i3.16030

Dekel B, Abrahams N. ‘I will rather be killed by corona than by him…’: Experiences of abused women seeking shelter during South Africa’s COVID-19 lockdown. López-Goñi JJ, ed. PLoS ONE. 2021;16(10):e0259275. doi:10.1371/journal.pone.0259275

Diaz A, Nucci-Sack A, Colon R, et al. Impact of COVID-19 Mitigation Measures on Inner-City Female Youth in New York City. Journal of Adolescent Health. 2022;70(2):220–227. doi:10.1016/j.jadohealth.2021.10.015

Ebert C, Steinert JI. Prevalence and risk factors of violence against women and children during COVID-19, Germany. Bull World Health Organ. 2021;99(6):429–438. doi:10.2471/BLT.20.270983

Egger D, Miguel E, Warren SS, et al. Falling living standards during the COVID-19 crisis: Quantitative evidence from nine developing countries. Sci Adv. 2021;7(6):eabe0997. doi:10.1126/sciadv.abe0997

El-Nimr NA, Mamdouh HM, Ramadan A, El Saeh HM, Shata ZN. Intimate partner violence among Arab women before and during the COVID-19 lockdown. J Egypt Public Health Assoc. 2021;96(1):15. doi:10.1186/s42506-021-00077-y

Every-Palmer S, Jenkins M, Gendall P, et al. Psychological distress, anxiety, family violence, suicidality, and wellbeing in New Zealand during the COVID-19 lockdown: A cross-sectional study. Francis JM, ed. PLoS ONE. 2020;15(11):e0241658. doi:10.1371/journal.pone.0241658

Gebrewahd GT, Gebremeskel GG, Tadesse DB. Intimate partner violence against reproductive age women during COVID-19 pandemic in northern Ethiopia 2020: a community-based cross-sectional study. Reprod Health. 2020;17(1):152. doi:10.1186/s12978-020-01002-w

Gibbons MA, Murphy TE, Rossi MA. Confinement and intimate partner violence. Kyklos. 2021;74(3):349–361. doi:10.1111/kykl.12275

Gresham AM, Peters BJ, Karantzas G, Cameron LD, Simpson JA. Examining associations between COVID-19 stressors, intimate partner violence, health, and health behaviors. Journal of Social and Personal Relationships. 2021;38(8):2291–2307. doi:10.1177/02654075211012098

Gulesci S, Puente–Beccar M, Ubfal D. Can youth empowerment programs reduce violence against girls during the COVID-19 pandemic? Journal of Development Economics. 2021;153:102716. doi:10.1016/j.jdeveco.2021.102716

Hamadani JD, Hasan MI, Baldi AJ, et al. Immediate impact of stay-at-home orders to control COVID-19 transmission on socioeconomic conditions, food insecurity, mental health, and intimate partner violence in Bangladeshi women and their families: an interrupted time series. The Lancet Global Health. 2020;8(11):e1380–e1389. doi:10.1016/S2214-109X(20)30366-1

Haq W, Raza SH, Mahmood T. The pandemic paradox: domestic violence and happiness of women. Peer J. 2020;8:e10472. doi:10.7717/peerj.10472

Hastuti L, Mardiani R, Rahmawati A, Wahyuni T, Kusumajaya S. Analysis of Risk Factors Related to the Events Domestic Violence (Qualitative Study on Women Victims of Violence During the Covid-19 Pandemic). International Journal of Progressive Sciences and Technologies. 2021;27(2):11. Available from: https://tinyurl.com/3n3vj6kh

Huq M, Das T, Devakumar D, Daruwalla N, Osrin D. Intersectional tension: a qualitative study of the effects of the COVID-19 response on survivors of violence against women in urban India. BMJ Open. 2021;11(9):e050381. doi:10.1136/bmjopen-2021-050381

Ibitoye TR, Ajagunna F. Sexual autonomy and violence against women in Nigeria: Assessing the impact of Covid-19 pandemic. De Jure. 2021;54. doi:10.17159/2225-7160/2021/v54a9

Indu PV, Vijayan B, Tharayil HM, Ayirolimeethal A, Vidyadharan V. Domestic violence and psychological problems in married women during COVID-19 pandemic and lockdown: A community-based survey. Asian Journal of Psychiatry. 2021;64:102812. doi:10.1016/j.ajp.2021.102812

Jetelina KK, Knell G, Molsberry RJ. Changes in intimate partner violence during the early stages of the COVID-19 pandemic in the USA. Inj Prev. 2021;27(1):93–97. doi:10.1136/injuryprev-2020-043831

Jung S, Kneer J, xKrüger THC. Mental Health, Sense of Coherence, and Interpersonal Violence during the COVID-19 Pandemic Lockdown in Germany. JCM. 2020;9(11):3708. doi:10.3390/jcm9113708

Karp C, Moreau C, Sheehy G, et al. Youth Relationships in the Era of COVID-19: A Mixed-Methods Study Among Adolescent Girls and Young Women in Kenya. Journal of Adolescent Health. 2021;69(5):754–761. doi:10.1016/j.jadohealth.2021.07.017

Lampe A, Daniels JK, Trawöger I, Beck T, Riedl D. Did domestic violence really increase in the early phase of the COVID-19 pandemic? Results of an interview-based observational study. Zeitschrift für Psychosomatische Medizin und Psychotherapie. 2021;67(3):303–314. doi:10.13109/zptm.2021.67.oa8

Lawson M, Piel MH, Simon M. Child Maltreatment during the COVID-19 Pandemic: Consequences of Parental Job Loss on Psychological and Physical Abuse Towards Children. Child Abuse & Neglect. 2020;110:104709. doi:10.1016/j.chiabu.2020.104709

Lee SJ, Ward KP, Rodriguez CM. Longitudinal Analysis of Short-term Changes in Relationship Conflict During COVID-19: A Risk and Resilience Perspective. J Interpers Violence. 2021 April:23. https://doi.org/10.1177/08862605211006359

Machlin L, Gruhn MA, Miller AB, et al. Predictors of family violence in North Carolina following initial COVID-19 stay-at-home orders. Child Abuse & Neglect. Published online October 2021:105376. doi:10.1016/j.chiabu.2021.105376

Maftei A, Dănilă O. Give me your password! What are you hiding? Associated factors of intimate partner violence through technological abuse. Curr Psychol. Published online August 10, 2021. doi:10.1007/s12144-021-02197-2

Mahapatro M, Prasad MM, Singh SP. Role of Social Support in Women facing Domestic Violence during Lockdown of Covid-19 while Cohabiting with the Abusers: Analysis of Cases Registered with the Family Counseling Centre, Alwar, India. Journal of Family Issues. 2021;42(11):2609–2624. doi:10.1177/0192513X20984496

Mahmood KI, Shabu SA, M-Amen KM, et al. The Impact of COVID-19 Related Lockdown on the Prevalence of Spousal Violence Against Women in Kurdistan Region of Iraq. J Interpers Violence. 2021 February. doi:10.1177/0886260521997929

Moawad AM, El Desouky ED, Salem MR, Elhawary AS, Hussein SM, Hassan FM. Violence and sociodemographic related factors among a sample of Egyptian women during the COVID-19 pandemic. Egypt J Forensic Sci. 2021;11(1):29. doi:10.1186/s41935-021-00243-5

Morgan A, Boxall H. Social Isolation, Time Spent at Home, Financial Stress and Domestic Violence during the COVID-19 Pandemic. Australian Institute of Criminology; 2020. doi:10.52922/ti04855

Moya A, Serneels P, Desrosiers A, Reyes V, Torres MJ, Lieberman A. The COVID-19 pandemic and maternal mental health in a fragile and conflict-affected setting in Tumaco, Colombia: a cohort study. The Lancet Global Health. 2021;9(8):e1068–e1076. doi:10.1016/S2214-109X(21)00217-5

Muldoon KA, Denize KM, Talarico R, et al. COVID-19 and perinatal intimate partner violence: a cross-sectional survey of pregnant and postpartum individuals in the early stages of the COVID-19 pandemic. BMJ Open. 2021;11(5):e049295. doi:10.1136/bmjopen-2021-049295

Naghizadeh S, Mirghafourvand M, Mohammadirad R. Domestic violence and its relationship with quality of life in pregnant women during the outbreak of COVID-19 disease. BMC Pregnancy Childbirth. 2021;21(1):88. doi:10.1186/s12884-021-03579-x

Oguntayo R, Oyeleke J, John-Oguntayo O, Aajayi-Hutchful F. Personality Traits, Emotional Intelligence, Socio-contextual Factors and Spousal Violence: The Trajectory of COVID-19 Pandemic Lockdown. IJBS. 2020;14(2). doi:10.30491/ijbs.2020.232959.1290

Ojeahere MI, Kumswa SK, Adiukwu F, Plang JP, Taiwo YF. Intimate Partner Violence and its Mental Health Implications Amid COVID-19 Lockdown: Findings Among Nigerian Couples. J Interpers Violence. Published online May 15, 2021:088626052110152. doi:10.1177/08862605211015213

Parrott DJ, Halmos MB, Stappenbeck CA, Moino K. Intimate partner aggression during the COVID-19 pandemic: Associations with stress and heavy drinking. Psychology of Violence. 2022;12(2):95–103. doi:10.1037/vio0000395

Pattojoshi A, Sidana A, Garg S, et al. Staying home is NOT ‘staying safe’: A rapid 8-day online survey on spousal violence against women during the COVID -19 lockdown in India. Psychiatry Clin Neurosci. 2021;75(2):64–66. doi:10.1111/pcn.13176

Phillimore J, Pertek S, Akyuz S, et al. “We are Forgotten”: Forced Migration, Sexual and Gender-Based Violence, and Coronavirus Disease-2019. Violence Against Women. Published online September 17, 2021:107780122110309. doi:10.1177/10778012211030943

Pinchoff J, Austrian K, Rajshekhar N, et al. Gendered economic, social and health effects of the COVID-19 pandemic and mitigation policies in Kenya: evidence from a prospective cohort survey in Nairobi informal settlements. BMJ Open. 2021;11(3):e042749. doi:10.1136/bmjopen-2020-042749

Plášilová L, Hůla M, Krejčová L, Klapilová K. The COVID-19 Pandemic and Intimate Partner Violence against Women in the Czech Republic: Incidence and Associated Factors. IJERPH. 2021;18(19):10502. doi:10.3390/ijerph181910502

Poonam, Pooja Tyagi. Correlates of domestic violence in relation to physical health and perceived stress during lockdown. Shodh Sarita. 2020;7(27):66–71. [cited 2022 Mar 1]. Available from: https://tinyurl.com/3v3zuaer

Raj A, Johns NE, Barker KM, Silverman JG. Time from COVID-19 shutdown, gender-based violence exposure, and mental health outcomes among a state representative sample of California residents. EClinicalMedicine. 2020;26:100520. doi:10.1016/j.eclinm.2020.100520

Rashid Soron T, Ashiq MAR, Al-Hakeem M, Chowdhury ZF, Uddin Ahmed H, Afrooz Chowdhury C. Domestic Violence and Mental Health During the COVID-19 Pandemic in Bangladesh. JMIR Form Res. 2021;5(9):e24624. doi:10.2196/24624

Rayhan I, Akter K. Prevalence and associated factors of intimate partner violence (IPV) against women in Bangladesh amid COVID-19 pandemic. Heliyon. 2021;7(3):e06619. doi:10.1016/j.heliyon.2021.e06619

Sabri B, Hartley M, Saha J, Murray S, Glass N, Campbell JC. Effect of COVID-19 pandemic on women’s health and safety: A study of immigrant survivors of intimate partner violence. Health Care for Women International. 2020;41(11-12):1294–1312. doi:10.1080/07399332.2020.1833012

Sari NP, van IJzendoorn MH, Jansen P, Bakermans-Kranenburg M, Riem MME. Higher Levels of Harsh Parenting During the COVID-19 Lockdown in the Netherlands. Child Maltreat. Published online June 17, 2021:107755952110247. doi:10.1177/10775595211024748

Schokkenbroek JM, Anrijs S, Ponnet K, Hardyns W. Locked Down Together: Determinants of Verbal Partner Violence During the COVID-19 Pandemic. Violence and Gender. 2021;8(3):148–153. doi:10.1089/vio.2020.0064

Sediri S, Zgueb Y, Ouanes S, et al. Women’s mental health: acute impact of COVID-19 pandemic on domestic violence. Arch Womens Ment Health. 2020;23(6):749–756. doi:10.1007/s00737-020-01082-4

Sharma P, Khokhar A. Domestic Violence and Coping Strategies Among Married Adults During Lockdown Due to Coronavirus Disease (COVID-19) Pandemic in India: A Cross-Sectional Study. Disaster med public health prep. Published online March 3, 2021:1-8. doi:10.1017/dmp.2021.59

Shokair zeinab, Abo Hamza E. Family Violence and its Impact on Children’s Mental Health During COVID-19 Pandemic. International Journal of Instructional Technology and Educational Studies. 2020;1(3):42–49. doi:10.21608/ihites.2020.42946.1035

Shrestha C, Ghimire C, Acharya S, Kc P, Singh S, Sharma P. Mental Wellbeing during the Lockdown Period following the COVID-19 Pandemic in Nepal: A Descriptive Cross-sectional Study. J Nepal Med Assoc. 2020;58(230). doi:10.31729/jnma.5498

Siegel A, Lahav Y. Emotion Regulation and Distress During the COVID-19 Pandemic: The Role of Childhood Abuse. J Interpers Violence. Published online June 4, 2021:088626052110219. doi:10.1177/08862605211021968

Spencer CM, Gimarc C, Durtschi J. COVID-19 Specific Risk Markers for Intimate Partner Violence Perpetration. J Fam Viol. Published online October 19, 2021. doi:10.1007/s10896-021-00335-9

Steinhoff A, Bechtiger L, Ribeaud D, et al. Self-Injury and Domestic Violence in Young Adults During the COVID-19 Pandemic: Trajectories, Precursors, and Correlates. J Res Adolesc. 2021;31(3):560–575. doi:10.1111/jora.12659

Tadesse AW, Tarekegn SM, Wagaw GB, Muluneh MD, Kassa AM. Prevalence and Associated Factors of Intimate Partner Violence Among Married Women During COVID-19 Pandemic Restrictions: A Community-Based Study. J Interpers Violence. Published online December 8, 2020:088626052097622. doi:10.1177/0886260520976222

Tesfaw LM, Kassie AB, Flatie BT. Sexual Violence and Other Complications of Corona Virus in Amhara Metropolitan Cities, Ethiopia. RMHP. 2021;Volume 14:3563–3573. doi:10.2147/RMHP.S297148

Teshome A, Gudu W, Bekele D, Asfaw M, Enyew R, Compton SD. Intimate partner violence among prenatal care attendees amidst the COVID-19 crisis: The incidence in Ethiopia. Int J Gynecol Obstet. 2021;153(1):45–50. doi:10.1002/ijgo.13566

Tierolf B, Geurts E, Steketee M. Domestic violence in families in the Netherlands during the coronavirus crisis: A mixed method study. Child Abuse & Neglect. 2021;116:104800. doi:10.1016/j.chiabu.2020.104800

Yamaoka Y, Hosozawa M, Sampei M, et al. Abusive and positive parenting behavior in Japan during the COVID-19 pandemic under the state of emergency. Child Abuse & Neglect. 2021;120:105212. doi:10.1016/j.chiabu.2021.105212

Yari A, Zahednezhad H, Gheshlagh RG, Kurdi A. Frequency and determinants of domestic violence against Iranian women during the COVID-19 pandemic: a national cross-sectional survey. BMC Public Health. 2021;21(1):1727. doi:10.1186/s12889-021-11791-9

Zhang H, Li Y, Shi R, Dong P, Wang W. Prevalence of Child Maltreatment during the COVID-19 Pandemic: A Cross-sectional Survey of Rural Hubei, China. The British Journal of Social Work. Published online August 25, 2021:bcab162. doi:10.1093/bjsw/bcab162

